# The Awareness of and Adherence to the Pregnancy Prevention Program for Oral Retinoids and Valproate: A Questionnaire Survey among Pharmacy Technicians Denmark

**DOI:** 10.64898/2026.05.13.26353084

**Authors:** Jasmin Hosseinzadeh, Ramune Jacobsen

## Abstract

**Background:** The use of oral retinoids and valproate during pregnancy can cause birth defects. In 2018, the EMA revised Pregnancy Prevention Programs (PPPs) for these medications. Pharmacy technicians in Denmark dispense prescription medications and must counsel customers.

**Aims:** This study aimed to examine knowledge of the teratogenicity of oral retinoids and valproate and use of the relevant PPPs among pharmacy technicians in Denmark.

**Methods:** A cross-sectional survey was conducted in spring 2025 using questionnaires developed for and tested in an international project. Data was collected via relevant Facebook groups and email invitations.

Descriptive statistics were used for analyses.

**Results:** For oral retinoids, 80 respondents were analyzed; 95% were women, 86% were pharmacy technicians, the mean age was 37.2 years. Most dispensed oral retinoids several times per month. Two respondents did not know retinoids were teratogenic. The most used PPP measure was the outer packaging warning (54%). Informing women about teratogenic effects was the most common practice. For valproate, 41 respondents were analyzed. Their characteristics were similar to those of respondents in the oral retinoid survey. Most dispensed valproate once per month. One-third did not know valproate was teratogenic. The outer packaging warning was used by 19%. The most common practice was referring to the prescribing physician if pregnancy was suspected.

**Conclusion:** Danish pharmacy technicians’ knowledge about teratogenic drugs and the PPP was poorer than that of pharmacists, especially regarding valproate, and requires attention in educational programs. The feasibility of PPP measures for both oral retinoids and valproate should be optimized.

## Introduction

Oral retinoids and valproate are widely used prescription medications indicated for severe dermatological and neurological disorders, respectively. Oral retinoids, such as isotretinoin, acitretin, and alitretinoin, are prescribed for severe acne and psoriasis (1). Valproate and related compounds, such as valproic acid, sodium valproate, magnesium valproate, valproate semi-sodium, and valpromide, are primarily used in the treatment of epilepsy and bipolar disorder and, in some countries, for migraine prophylaxis (2). Alongside their therapeutic benefits, both medication classes are strongly associated with substantial teratogenic risks. Prenatal exposure to oral retinoids has been reported to result in congenital malformations and long-term neurodevelopmental impairments in approximately 25% of exposed infants (3). Valproate exposure during pregnancy has been linked to major congenital malformations and developmental disorders in around 10% of prenatally exposed individuals (4, 5).

In the context of safety communication and in response to persistent prenatal exposure to oral retinoids and valproate, the European Medicines Agency (EMA) revised Pregnancy Prevention Programs (PPPs) in 2018 for both medications (6, 7). The programs were designed to ensure that women of childbearing potential are informed of teratogenic risks and protected from unintended pregnancies while receiving treatment. The revised PPPs included comprehensive risk minimization measures such as educational materials for healthcare professionals and patients, direct healthcare professional communication letters (DHPCs), patient reminder cards, structured risk acknowledgement forms, pharmacy checklist, and information within and outside medication packaging.

To evaluate awareness of and adherence to the revised PPPs, the EMA supported a multi-country surveys conducted in 2019–2020 across eight European countries: Belgium, Denmark, Greece, Latvia, Portugal, the Netherlands, Slovenia, and Spain (8, 9). The surveys aimed to assess where physicians, pharmacists, and patients had obtained information about teratogenic risks associated with oral retinoids and valproate, and the extent to which they adhered to the PPPs. Findings revealed that despite general awareness of the teratogenicity of these medications, the utilization of PPP materials and risk minimization tools remained limited across all stakeholder groups. Importantly, pharmacists in Denmark exhibited comparatively lower levels of knowledge about the teratogenic risks of valproate relative to pharmacists in other participating countries (10).

In Denmark, pharmacy technicians constitute a key professional group responsible for dispensing prescription medications and providing counselling to customers in community pharmacies (11, 12). In routine practice, they frequently interact with women of childbearing potential who may be receiving teratogenic medications such as oral retinoids and valproate. Despite their critical frontline role, research examining pharmacy technicians’ knowledge, counselling practices, or their engagement with PPP tools is scarce. To date, no studies have explored how Danish pharmacy technicians manage high-risk medicines or how they support risk minimization during the dispensing of teratogenic medications. Therefore, the aim of the present study was to investigate Danish pharmacy technicians’ knowledge regarding the teratogenicity of oral retinoids and valproate, as well as to explore the use of the revised PPPs when counselling women of childbearing age who purchase these medications in Danish community pharmacies.

## Methods

### Study design and instruments

Two cross-sectional questionnaire surveys were conducted in March–May 2025 using questionnaires developed for an international survey and previously tested among Danish pharmacists (8, 9). The questionnaire for pharmacy technicians inquired about: 1) their awareness of, and information sources regarding, the teratogenic effects of oral retinoids and valproate; 2) their knowledge of and adherence to the PPP for the respective medication; 3) their practices when dispensing these medications; and 4) whether the dispensing practices changed in 2018 when the EMA updated the PPPs. Additionally, in both surveys an open-ended question was included, asking respondents to list the barriers for using the PPP’s measures. The questionnaires in English can be found in the publicly available reports of the international studies (8, 9).

### Sampling approach and procedures

Data was collected online using SurveyXact system (13). Links to the questionnaire were provided in the relevant Facebook groups. Additionally, emails with an invitation to participate were sent to all members of the Danish Pharmacy Practice Research Network (14) and to every second pharmacy e-mail listed on the Danish medicine information site (15). A reminder was sent after one week.

### Data analysis

Descriptive analyses were performed for categorical variables using frequencies and percentages. SPSS version 29 was used for all statistical analyses and Microsoft Excel was utilized for data visualization. Answers to the open-ended questions were examined through qualitative content analysis.

### Informed Consent

All participants provided informed consent before responding to the questionnaire. The data were collected in anonymized form.

## Results

### Sample characteristics

Table 1 presents the characteristics of the respondents in the oral retinoid and valproate studies. In total, 80 respondents were included in the oral retinoid study. Of these, 95% were women, 86% were pharmacy technicians, and 15% were pharmacy technician students. The mean age was 37.2 years. Most had worked in a pharmacy for 0–5 years and dispensed oral retinoids to women of reproductive age several times per month. The number of included respondents in the valproate study was almost half that: 41. The distribution of their gender, age, profession and work experience categories was similar to those of the respondents in the oral retinoid study, except that most dispensed valproate only once per month.

**Table 1.** Characteristics of participants.

|  |  | **Ora retinoids**<br>**survey N=80** | **Valproate survey**<br>**N=41** |
| --- | --- | --- | --- |
| **Age** | Mean (SD) | 37.3 (12.16) | 33.1 (9.21) |
|  | Median (IQR) | 33.00 (28.00-46.75) | 29 (26.5-41.0) |
|  | Range | 19-69 | 21-58 |
| **Gender, N (%)** | Female | 76 (95.0%) | 37 (90.2%) |
|  | Male | 2 (2.5%) | 2 (4.9%) |
|  | Rather not say | 2 (2.5%) | 2 (4.9%) |
| **Professional category,**<br>**N (%)** | Pharmacy technician | 68 (85.0%) | 33 (80.5%) |
|  | Student pharmacy technician | 12 (15.0%) | 8 (19.5%) |
| **Period practicing**<br>**current profession, N**<br>**(%)** | 0-5 years | 43 (53.8%) | 25 (61.0%) |
|  | 6-10 years | 13 (16.3%) | 7 (17.1%) |
|  | 11-20 years | 15 (18.8%) | 3 (7.3%) |
|  | Over 20 years | 9 (11.3%) | 6 (14.7%) |
| **Frequency of**<br>**dispensing oral**<br>**retinoids for women of**<br>**reproductive age, N (%)** | Once a week or more | 22 (27.5%) | 3 (7.3%) |
|  | A few times a month | 35 (43.8%) | 10 (24.4%) |
|  | Once a month or less frequently | 23 (28.7%) | 28 (68.3%) |

### Teratogenicity awareness and information sources

Results on respondents’ awareness of the teratogenicity of the two medications, and the information sources, are shown in Table 2. In the oral retinoid study, two participants (2.5%) were not aware that oral retinoids are teratogenic; the remaining respondents mainly obtained their knowledge from studies (54%) and colleagues (39%). In the valproate study, nearly one-third of participants were unaware of the teratogenicity; as with oral retinoids, the remainder primarily obtained their knowledge from studies (44%) and colleagues (22%).

**Table 2:** Awareness about the teratogenic effect of oral retinoids and valproate.

|  |  | **Oral retinoids**<br>**study, N = 80** | **Valproate**<br>**study, N = 41** |
| --- | --- | --- | --- |
| **When first**<br>**learned about the**<br>**teratogenic effect** | Within the last 2 years | 17 (21.3%) | 6 (14.6%) |
|  | Withing the last 5 years | 17 (21.3%) | 12 (29.3%) |
|  | More than 5 years ago | 44 (55.0%) | 11 (26.8%) |
|  | Just now | 2 (2.5%) | 12 (29.3%) |
| **Where from first**<br>**learned about the**<br>**teratogenic effect** | Health Authorities | 16 (20.0%) | 4 (9.8%) |
|  | Danish Medicines Agency | 20 (25%) | 6 (14.6%) |
|  | Professional Societies | 6 (7.5%) | 2 (4.9%) |
|  | Colleagues | 31 (38.8%) | 9 (22.0%) |
|  | Professional Journals | 4 (5.0%) | 4 (9.8%) |
|  | Manufacturers | 19 (23.8%) | 6 (14.6%) |
|  | Internet | 8 (10.0%) | 3 (7.3%) |
|  | Symposia/conferences | 0 (0.0%) | 1 (2.4%) |
|  | Academic/professional studies | 43 (53.8%) | 18 (43.9%) |
|  | Post-academic training | 4 (5.0%) | 1 (2.4%) |
|  | Other | 8 (10.0%) | 1 (2.4%) |

### Patterns of the use of materials from the PPPs

Figure 1ab, show the patterns of current and potential future use of the different PPP materials for both medications. In both cases, the warning label on the outer packaging was the most frequently used material. Among participants who did not currently use the materials and were asked whether they would consider using them in the future, different patterns were observed for different medications. For oral retinoids, except for the outer packaging warning, more respondents did not intend than intend to use the materials in the future (Figure 1a). For valproate, in opposite, more intended than not intended to use the materials in the future (Figure 1b).

**Figure 1a.**
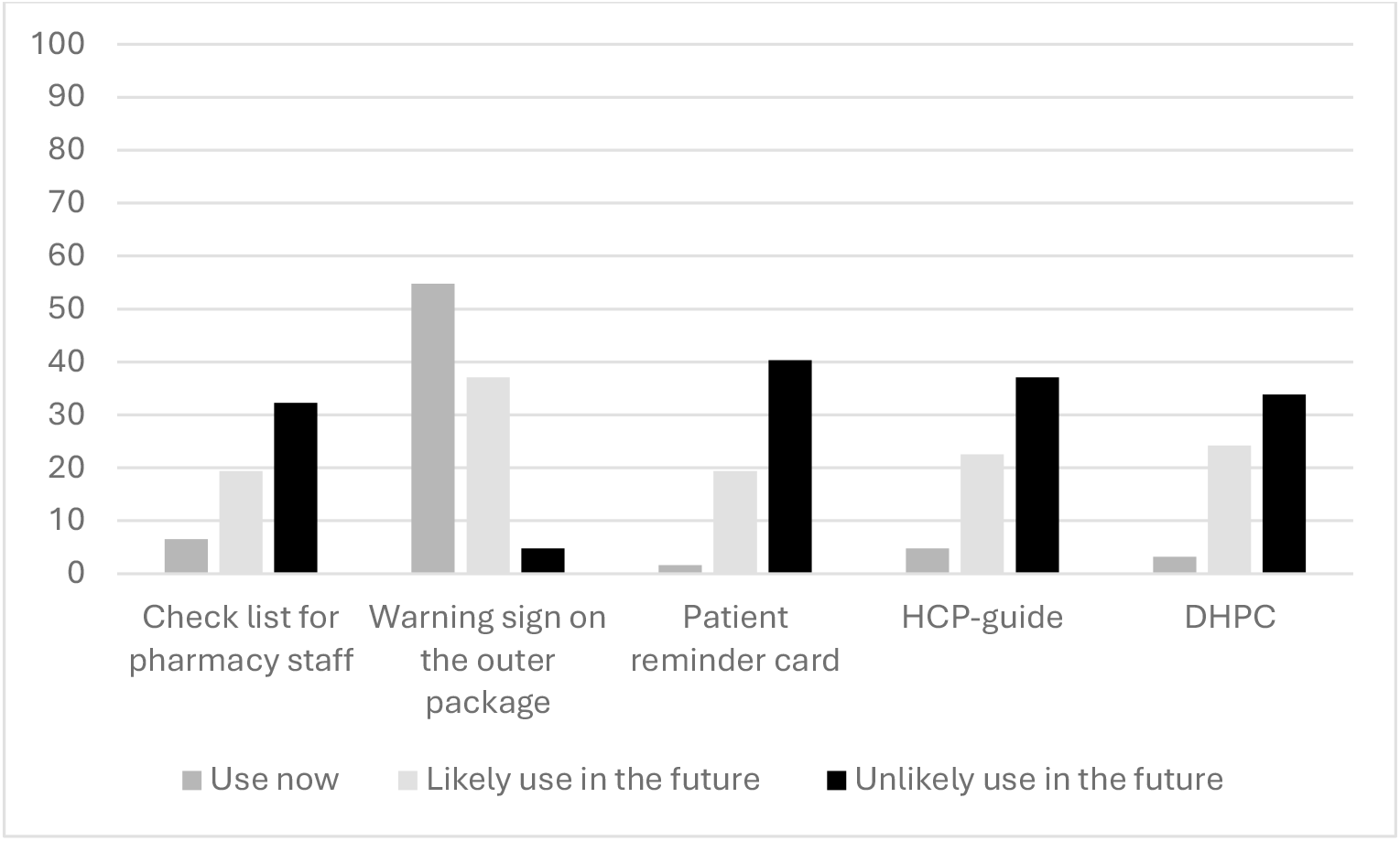
Patterns of the use of materials from PPP for oral retinoids, %

**Figure 1b.**
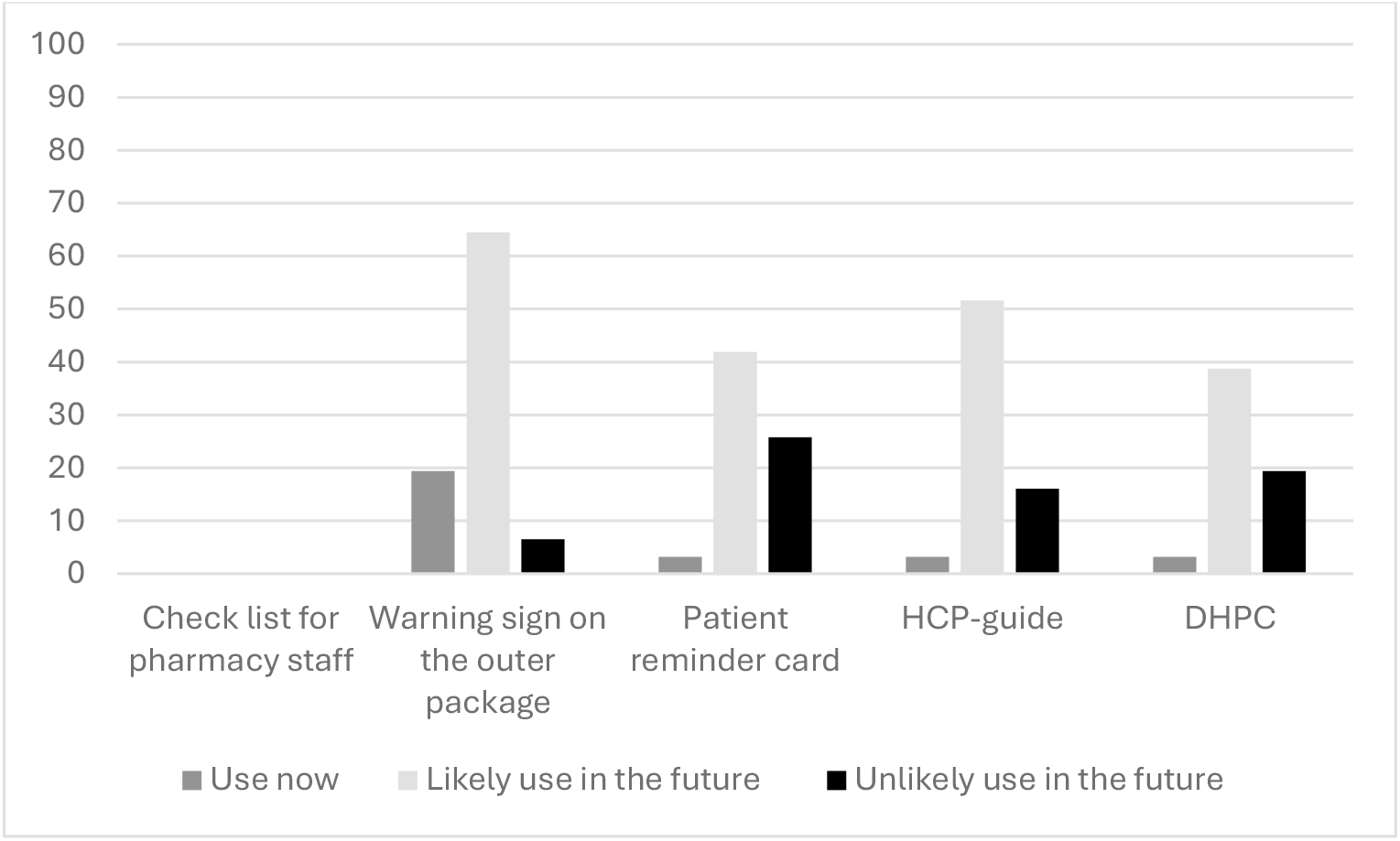
Patterns of the use of materials from PPP for valproate, %

### Dispensing practices and their changes after the PPP update

The most common practice when dispensing oral retinoids was informing women about teratogenic effects (72%); the most common practice when dispensing valproate was referring to the prescribing physician if pregnancy was suspected (62%), followed by informing women about teratogenic effects (45%) (Supplementary material, table S2). In total, nine respondents in the oral retinoid study and eight in the valproate study reported that their practice regarding the dispensing of teratogenic medication changed in 2018, when the EMA updated the PPP. For both medications, the most impactful PPP measure associated with this change was the warning on the outer packaging (Supplementary material, Table S3).

### Barriers for using PPP measures

Qualitative content analysis of the open-ended question on barriers to implementing the PPP in daily pharmacy practice when dispensing teratogenic medicines identified several overarching themes that were common to both oral retinoids and valproate: lack of knowledge about PPP materials; non practical design of the materials; time pressure in routine pharmacy work; the perception that responsibility for communicating teratogenic risk rests with the prescribing physician; and the perception that customers do not wish to discuss the topic due to its sensitivity (i.e., pregnancy). For valproate, an additional theme emerged: because the medication is dispensed so rarely, participants reported forgetting about the PPP. Table 3 presents key quotations exemplifying these themes in the oral retinoid and valproate studies.

**Table 3.** Barriers for the use of PPP and examples of quotations from the open ended-question.

| Themes | Quotations from the oral retinoid survey | Quotations from the valproate survey |
| --- | --- | --- |
| **Lack of knowledge** | *“Have not seen the materials”*<br>*“Lack of knowledge about these things”* | *“Too little information, didn’t know there was a guideline”*<br>*“Have never seen the healthcare professionals’ guideline, patient card, letter”* |
| **Impracticality** | *“All the material that is produced is too complicated and makes it unmanageable for the pharmacy staff”* | *“Paper is not effective today”*<br>*“Cards get lost”* |
| **Time pressure** | *“Busyness/time”*<br>*“Time”*<br>*“The busyness”* | *“Time”*<br>*“Must prioritize the most important first”* |
| **Physicians’ responsibility** | *“It is the responsibility of the prescribing doctor.”*<br>*“They [patients/customers] are often very well informed by the specialist”* | *“Expect the doctor to take responsibility”*<br>*“One assumes that the doctor has informed the patient/customer”* |
| **Sensitivity of the topic** | *“Personal topic”*<br>*“Modesty/embarrassment when talking about pregnancy with young girls”*<br>*“Most often they [patients/customers] do not want to talk about it”* | *“Very sensitive topic”*<br>*“Delicate topic”*<br>*“Not discreet”* |
| **Forgetfulness** |  | *“So rare that we dispense Valproate. The information gets forgotten”* |

## Discussion

The study investigated Danish pharmacy technicians’ awareness of the teratogenicity of oral retinoids and valproate, as well as their adherence to EMA-issued educational materials and prevention measures while dispensing these medications to women of childbearing age, updated in 2018. The study showed that Danish pharmacy technicians were generally aware of the teratogenicity of oral retinoids but less aware of that of valproate, relying mainly on studies and colleagues for information. Oter packaging warnings were the most used PPP measures for both medications, though intentions to use PPP’s measures in the future differed for the two medications. While dispensing the medications, study participants most often informed women about teratogenic risks for oral retinoids and referred suspected pregnancies to physicians for valproate. Few adjusted their behavior after the PPPs’ update in 2018. Barriers to using PPP measures included limited knowledge of the materials, their impractical design, time pressure, and some attitudes discouraging the use.

### Awareness of teratogenicity and information sources

The study revealed substantial variability in pharmacy technicians’ awareness of teratogenic risks, with notably lower knowledge levels regarding valproate compared with oral retinoids. While awareness of the teratogenic risks associated with oral retinoids was relatively high, nearly one-third of respondents were not aware of teratogenic risks of valproate. When comparing these results with the data from the previously published surveys among Danish pharmacists, pharmacy technicians demonstrated awareness levels for oral retinoids similar to those of pharmacists (16), but exhibited substantially lower awareness regarding valproate (10). Several factors may explain this discrepancy. First, oral retinoids have been the focus of regulatory attention and risk minimization activities for a much longer period. Risk minimization measures for oral retinoids were introduced as early as 2003, whereas structured risk minimization activities for valproate were implemented approximately a decade later, in 2014 (6, 7) Over time, awareness of the teratogenicity of oral retinoids thus had received greater attention. Second, oral retinoids are being prescribed more frequently and to younger women of childbearing potential (10, 16), which makes awareness of their teratogenicity more urgent

Regarding the differences in teratogenicity awareness between pharmacy technicians and pharmacists, one explanation may be the education programs of the two professional groups in Demark. Pharmacist education is longer (five years, compared with three years for pharmacy technicians) and includes more theoretical training, whereas pharmacy technicians acquire most of their knowledge through practice-based learning (17). Moreover, the previous and this study found that the two professions tended to rely on different information sources. Pharmacy technicians more often reported relying on colleagues, whereas pharmacists more often obtained information from postgraduate education (10, 16). This may reflect differences in access to, or engagement with, continuing education, potentially leaving technicians less consistently exposed to safety updates from the authorities. Findings from the study examining Danish pharmacists’ and pharmacy technicians’ knowledge differences in other pharmaceutical area support this interpretation (18).

In summary, these findings point to a potentially problematic situation. In Denmark, pharmacy technicians are responsible for dispensing medicines as often as pharmacists, and hence, they play an important role in customer counseling. Pharmacy technicians’ insufficient baseline knowledge about serious risks, such as the teratogenicity of valproate, can have implications for patient safety. The observed gaps therefore merit attention from a pharmacy technicians’ education perspective to ensure that all pharmacy staff involved in dispensing high-risk medicines are timely updated.

### Actual and future use of PPPs materials

This study identified two contrasting patterns in pharmacy technicians’ engagement with PPP materials. The first pattern was observed for valproate, where current use of PPP materials was limited, but there was a higher wish to use the materials in the future. This pattern suggests that the low adherence to the valproate’s PPP may be driven by a lack of awareness: many participants were unfamiliar with the materials prior to the study, but once introduced, most expressed a clear willingness to use them in the future. The pattern thus further highlights the need for educational interventions, such as better dissemination of information or brief training activities about the requirements of the PPP for valproate’.

The second pattern concerned oral retinoids, where awareness of teratogenic risk was relatively high but willingness to adopt PPP tools in the future was low. This suggests that non-use is not caused by lack of knowledge but rather by some attitudinal barriers, for example, perceived lack of added value of the PPP for oral retinoids, difficulties of incorporating the use of the PPP into daily workflow, or unclarity of the medication dispenser role. The limited engagement with the pharmacist checklist, despite its being a central PPP component for pharmacy staff, is particularly notable. This pattern thus points to the need for more nuanced qualitative exploration of how pharmacy technicians evaluate their role, and how they view the usability and necessity of different PPP elements in everyday dispensing situations.

When compared with previously reported data from the pharmacists’ survey, pharmacy technicians were less likely to currently use PPP materials but reported a stronger intention to use them in the future, particularly for valproate (10, 16). This further supports the insight that for valproate, the main barriers to using PPP measures are lack of teratogenicity awareness and lack of knowledge about the PPP materials.

In summary, the different patterns illustrate that effective implementation of PPP materials may depend on the product involved. For valproate, increasing awareness appears to be the key. For retinoids, improving usability, clarifying professional responsibilities, and ensuring better workflow to integrate risk minimization measures seem to be more important. To address the gaps in PPP implementation in pharmacies, these product-related differences should be taken into account.

### Dispensing practices

In line with the different patterns of adhering to the PPPs for oral retinoids and valproate, pharmacy technicians’ reported dispensing practices for different medications were also different. For example, for oral retinoids, 72.4% of pharmacy technicians stated that they regularly informed patients about contraception, while only 44.8% did so when dispensing valproate. When comparing with pharmacists, pharmacy technicians reported “always” and “often” performing the selected practices less frequently than pharmacists when dispensing oral retinoids (16). However, when dispensing valproate, across the same set of practices, pharmacy technicians selected “always” or “often” more frequently than pharmacists (10). This divergence suggests that the level of engagement in risk-mitigation activities vary not only by medication type but also by profession. For oral retinoids, where pharmacy technicians seemed to be aware about teratogenic risks on a level comparable to that of pharmacists, the counselling practices were performed less frequently, possibly due to uncertainty regarding the technician’s role. For valproate, where teratogenicity awareness among all the pharmacy technicians was low, those who were knowledgeable - likely a selective subgroup – reported providing counselling at a level similar to that of pharmacists.

The generalizability of the results regarding changes in dispensing practices for both oral retinoids and valproate is uncertain due to the small sample sizes. Regardless, the study showed that most pharmacy technicians were unsure whether their dispensing practices had changed following the implementation of the PPPs in 2018 for both medications. Among those who did report that changes happened, the warning label on the outer packaging was the most frequently reported material influencing the change. This finding is consistent with trends observed among pharmacists, where visual measures, i.e. warning labels on the outer package, were more likely to be noticed and acted upon than formal PPP materials, e.g., pharmacy staff checklist (10, 16). Consistent with the barriers to PPP use reported in this study, this underscores that the practicality of the measures, such as ease of access or compatibility with daily workflow, is just as important as awareness to ensure the application of the measures in routine pharmacy practice.

### Strengths and Limitations

This was one of the few studies exploring dispensing practices among Danish pharmacy technicians, and the first one focusing on awareness of teratogenicity and dispensing practices related to high-risk medications such as oral retinoids and valproate. A strength of the study was the use of questionnaires that had been developed and tested among pharmacies in an international study. The main limitation was the small sample size, which limits the generalizability of the findings. The sample size was particularly low for the valproate study, indicating a perceived low relevance of the topic (i.e. dispensing of valproate to women of childbearing potential is a rare occasion) and the self-selection of the most motivated respondents. This could further indicate that the awareness of valproate teratogenicity and the use of the related PPP is underestimated in the study. That is, the self-selection of the most interested participants could potentially have led to an overrepresentation of pharmacy technicians with a personal or professional interest in the topic and with greater knowledge than average. Therefore, the indicated knowledge gaps concerning the teratogenicity of valproate and the need for targeted educational interventions may, in fact, be even more pronounced than reflected in our findings.

## Conclusion

Pharmacy technicians’ knowledge about the teratogenicity of oral retinoids and valproate was poorer than that of pharmacists, especially in relation to valproate. Their use of measures from the PPPs for oral retinoids was similar to that of pharmacists, but use of the measures from the PPP for valproate was markedly lower. This indicates a need for greater emphasis on educating pharmacy technicians about teratogenic medicines and the PPPs, particularly regarding valproate. Moreover, the feasibility of PPP measures for both oral retinoids and valproate should be optimized.

## Data Availability

The data that support the findings of this study are available from the corresponding author upon reasonable request.

## Acknowledgements

None

## Ethics declaration

All participants provided informed consent before responding to the questionnaire. The data were collected in anonymized form. According to Danish law, a formal ethical assessment was not necessary, as the study did not collect any biological material.

## Conflict of Interest Statement

The authors declare they have no conflicts of interest

## Funding Statement

This research received no specific grant from any funding agency in the public, commercial, or not-for-profit sectors.

## Open Access Statement

No specific open access funding received.

## Notes

### Competing Interest Statement

The authors have declared no competing interest.

### Funding Statement

This study did not receive any funding

