## Supplementary material for "The Awareness of and Adherence to the Pregnancy Prevention Program for Oral Retinoids and Valproate: A Questionnaire Survey among Pharmacy Technicians Denmark"

**Table S1a.** The use of education materials from the PPP, N (%)

|  | **Oral retinoids study,** N=62 | **Valproate study,** N=31 |
| --- | --- | --- |
| **Check list for pharmacy staff** | 4 (6.5%) | n.a. |
| **Warning sign on the outer package** | 34 (54.8%) | 6 (19.4%) |
| **Patient reminder card** | 1 (1.6%) | 1 (3.2%) |
| **HCP-guide** | 3 (4.8%) | 1 (3.2%) |
| **DHPC** | 2 (3.2%) | 1 (3.2%) |

**Table S1b.** The use of education materials from the PPP in the future among those who do not use them, N (%)

|  |  | **Oral retinoids study,** N=62 | **Valproate study,** N=31 |
| --- | --- | --- | --- |
| **Check list for pharmacy staff** | Likely to use | 12 (19.4%) | n.a. |
|  | Unlikely to use | 20 (32.3%) | n.a. |
| **Warning sign on the outer package** | Likely to use | 23 (37.1%) | 20 (64.5%) |
|  | Unlikely to use | 3 (4.8%) | 2 (6.5%) |
| **Patient reminder card** | Likely to use | 12 (19.4%) | 13 (41.9%) |
|  | Unlikely to use | 25 (40.3%) | 8 (25.8%) |
| **HCP-guide** | Likely to use | 14 (22.6%) | 16 (51.6%) |
|  | Unlikely to use | 23 (37.1%) | 5 (16.1%) |
| **DHPC** | Likely to use | 15 (24.2%) | 12 (38.7%) |
|  | Unlikely to use | 21 (33.9%) | 6 (19.4) |

Table S2. Practices when dispensing teratogenic medications to women of reproductive age

|  | **Oral retinoids study**, N=58 | **Valproate study, N=29** |
| --- | --- | --- |
| Inform about effective contraception | 42 (72.4%) | 13 (44.8%) |
| Advice stopping treatment when pregnant | 21 (36.2%) | 21 (72.4%) |
| Advice to contact doctor when suspect a pregnancy | 33 (56.9%) | 11 (37.9%) |
| Inform about pregnancy testing before/ during/after treatment | 31 (53.4%) | 20 (69.0%) |

**Table S3**. If change occurred and which PPPs changed practices of dispensing teratogenic medications

|  |  | **Oral retinoids study**, N = 54 | **Valproate study**, N = 26 |
| --- | --- | --- | --- |
| **If change occurred** | Yes | 9 (16.7%) | 8 (30.7%) |
|  | Not sure | 27 (50.0%) | 11 (42.3%) |
|  | No | 18 (33.4%) | 7 (26.9%) |
| **Which PPP changed the practices** | Check list for pharmacy staff | 2 (22.2% from 9) | n.a. |
|  | Warning sign on the outer package | 8 (88.9% form 9 | 6 (75.0% from 8) |
|  | Patient reminder card | 1 (11.1% fro 9) | 2 (25.0% from 8) |
|  | HCP-guide | 2 (22.2% from 9) | 3 (37.5% from 8) |
|  | DHPC | 1 (11.1% from 9) | 2 (25.0% from 8) |
